# Time series analysis of routine immunisation coverage during the COVID-19 pandemic in 2021 shows continued global decline and increases in Zero Dose children

**DOI:** 10.1101/2023.02.06.23285411

**Authors:** Beth Evans, Olivia Keiser, Laurent Kaiser, Thibaut Jombart

## Abstract

Whilst it is now widely recognised that routine immunisation (RI) was disrupted by the COVID-19 pandemic in 2020 compared to previous immunisation performance, the extent of continued interruptions in 2021 and/or rebounds to previous trends remains unclear, with sporadic surveys reporting signs of immunisation system recovery at the end of 2020.

We modelled country-specific RI trends using validated estimates of national coverage from the World Health Organisation and United Nation Children’s Fund for over 160 countries, to project expected diphtheria, tetanus, and pertussis-containing vaccine first-dose (DTP1), third-dose (DTP3) and measles-containing vaccine first-dose (MCV1) coverage for 2021 based on pre-pandemic trends (from 2000-2019).

We estimated a 3·6% (95%CI: [2·6%; 4·6%]) decline in global DTP3 coverage in 2021 compared to 2000-2019 trends, from an expected 90·1% to a reported 86·5% across 164 reporting countries, and similar results for DTP1 (2·8% decline; 95%CI: [2·0%; 3·6%]), and for MCV1 (3·8% decline; 95%CI: [4·8%; 2·7%]). 86·5% global coverage in 2021 represents a further decrease from that reported in 2020 and 2019, and translates to a 16-year setback in RI coverage, i.e., 2005 levels. Hypothesised and early signals of rebounds to pre-pandemic coverage were not seen in most countries. The Americas, Africa, and Asia were the most impacted regions, with low- and middle-income countries the most affected income groups.

The number of Zero Dose children also continued to increase in 2021. DTP1 coverage declined worldwide from an expected 93·7% to a reported 90·9% (2·8% decline; 95%CI: [2·0%; 3·6%]) which translates into an additional 3.4 million Zero Dose children on top of an expected 11.0 million (30.9% increase) at the global level.

We hope this work will provide an objective baseline to inform future interventions and prioritisation aiming to facilitate rebounds in coverage to previous levels and catch-up of growing populations of under- and un-immunised children.

## Introduction

It is now widely recognised that routine immunisation (RI) was disrupted globally during the start of the COVID-19 pandemic and throughout 2020 [1–4]. Single-country studies, and literature reviews have reported delays and interruptions in service delivery, decreases in attendance, and/or full suspension of services during the pandemic, e.g., during periods of quarantine or implementation other non-pharmaceutical interventions. Observational and modelling studies have evaluated the impact of disruptions at the country-, regional-, and global-level in terms of coverage reduction, with estimates ranging from 2% [5] to over 7% globally in 2020 [6]. We previously published an estimated 2.9% decline for diphtheria, tetanus, and pertussis-containing vaccine third-dose (DTP3; 95% CI: 2.2%; 3.6%) and similar declines for other key vaccine indicators [7]. Prior research has established associations between lower vaccination coverage and outbreaks of vaccine preventable diseases (e.g., Measles outbreaks in Zambia [8] and the United States of America [9]), highlighting the potential health implications of the knock-on effects of the pandemic.

Whilst the evidence of RI disruptions in 2020 is now well documented, it remains unclear whether such disruptions continued in 2021. Few papers have been published on 2021 coverage, with some exceptions largely focussing on single countries (e.g., Brazil – [10]), and the grey literature hinting at a potential decline in global coverage between 2020 and 2021 [11,12].

Here, we build on previously published methods to test and quantify the extent of RI disruption in 2021 compared to pre-pandemic trends. Reaching Zero Dose children (those that receive no vaccinations) is crucial to reduce the most acute vaccine-preventable morbidity and mortality in infants. Therefore, we translate modelled coverage disruptions into quantification of under- and un-immunised (i.e., Zero Dose) children. We also investigate year-on-year trends within countries to identify rebounds and continued declines, with the aim to facilitate future investigations of the key factors underpinning these dynamics and focus efforts and resources to aid recovery.

## Materials and methods

### Data collection

Building on our previously published methodology to identify deviations from temporal trends in RI [7], we investigated changes in RI coverage using three key indicators: diphtheria, tetanus, and pertussis-containing vaccine first-dose (DTP1) and third-dose (DTP3) coverage, and measles-containing vaccine first-dose (MCV1). To recap – DTP1 is typically administered at around six-weeks of age and is used as a proxy for inequity to quantify Zero Dose children [13]; DTP3 is delivered to children of approximately fourteen-weeks old and serves as a general marker for immunisation system performance, used by national and global immunisation stakeholders; and MCV1 is recommended to be delivered at nine-months of age and is often used as an additional indicator of health system performance.

We used coverage data published by the World Health Organisation and United Nation Children’s Fund (UNICEF) Estimates of National Immunisation Coverage (WUENIC [14,15]) from 2000-2021 inclusive, using the latest (July 2022) WUENIC data release [16]. It is noted that included within the latest WUENIC release are retrospective revisions to previous-year (*i*.*e*., pre-2021) coverage estimates for the Democratic Republic of the Congo, Samoa, Tunisia, Guatemala, Philippines, and Cote d’Ivoire [16]. Such revisions typically represent validation of population-level coverage surveys that replace previously reported coverage levels for the current and some recent years. For this reason, we have updated predictive modelling for expected coverage in 2020 using the latest WUENIC data rather than leveraging previous results [7].

Population data was sourced from the United Nations World Population Prospects 2022 [17] release (UNWPP 2022, including retrospective updates of 2021 populations). Income group classification was taken from the World Bank’s 2021 categorisation of countries [18].

### Statistical analysis

We used Auto Regressive Integrated Moving Average (ARIMA) modelling [19] to capture temporal trends in coverage for each country from 2000-2019 to forecast expected coverage in 2020 and 2021. The inclusion of 2020 in the ARIMA model fitting to forecast coverage in 2021 was ruled out as many countries showed sharp changes in point estimates of coverage that year. While this may result in wider confidence intervals (CIs) for 2021 than 2020, results were essentially unchanged when also including 2020 in the model-fitting stage for countries which did not show sharp changes in 2020.

Prior to investigating global, regional, income group, and year-on-year differences between expected and observed coverage, historic (pre-2020) and predicted time series were assessed, and countries were removed from analyses for one of two reasons:

#### Prior knowledge

Countries lacking recent WUENIC coverage updates: For 7 countries (Cambodia, the Central African Republic, Haiti, Guinea, Lesotho, Somalia, and South Sudan), WUENIC report an inability to update coverage estimates for successive years due to the absence of additional information meeting their criteria, resulting in WUENIC coverage estimates appearing as constant. In addition, countries with large geopolitical events known to have affected immunisation were removed. This applied to Myanmar and the Democratic People’s Republic of Korea, where immunisation coverage reportedly fell over 60%between 2020 and 2021. Whilst some of these coverage declines may have been associated with the pandemic, to be conservative we removed these countries for aggregate analyses.

#### Poor model fit

Poor model fit was identified in countries where projected coverage in 2020 fell outside either two standard deviations or six percentage points from WUENIC-reported 2019 coverage. Both of these scenarios were deemed to indicate programmatically unlikely changes in coverage (either infeasible programmatic improvements or major disruptions). These countries were removed to be conservative and not overly associate changes in coverage with pandemic impact, where the explanation may be driven by poor model fitting (e.g., due to prior year volatility, or skew from anomalous data points).

The list of countries, per vaccine dose, that were removed due to lack of reliability in assessing temporal trends, are detailed in the Appendix (Section S4). We conducted a sensitivity analysis on aggregate, regional and income group level analyses with no countries excluded – these are detailed in the Appendix (Section S7).

Changes in coverage were measured as the difference between the reported and expected coverage for a given year. Values are reported as percentages and these refer to percentage point changes. The following analyses were completed for each vaccine dose:

#### Comparing expected versus reported coverage for 2021

We conducted *t-tests* of reported vaccination coverage against the null hypothesis of the absence of change in trends from prior years for 2021 at the global level, and by region and income group – to investigate the existence of RI impact in 2021. We then investigate regional and income group-level heterogeneities, using linear models with coverage changes as a response variable and ANOVA using the corresponding categorisation of countries. Differences between individual countries were assessed by comparing the 95% CIs of the linear models’ coefficients.

#### Calculating additional missed immunisations

We calculated the additional number of missed immunisations by combining the estimated changes in coverage (including estimates of low- and high-bounds using 95% CIs) with surviving infant population estimates reported in the UNWPP 2022 dataset. These results indicate the number of additional unvaccinated children per country that would have been reached based on previous immunisation trends.

#### Trends in coverage from 2020 to 2021

We investigated absolute changes in coverage between 2020 and 2021 per country to quantify the extent of recovery and continued coverage declines. We tested whether trends in 2020 were associated with trends in 2021, by conducting Fisher’s Exact tests on the relevant contingency table.

All analyses were conducted using R, version 4.1.2 [20] and can be reproduced using a publicly available *reportfactory* including required data (all publicly available) and scripts [21].

## Results

### 2021 coverage

After removing countries further analyses were conducted on: 164 countries for DTP3, 168 countries for DTP1, and 160 countries for MCV1. Examples of coverage trends for the largest countries included are shown in **Figure 1** for each vaccine dose. Similar trends can be seen within countries across different vaccine doses (with DTP1 and DTP3 being more similar than MCV1 coverage trends), and large differences between countries.

**Figure 1.**
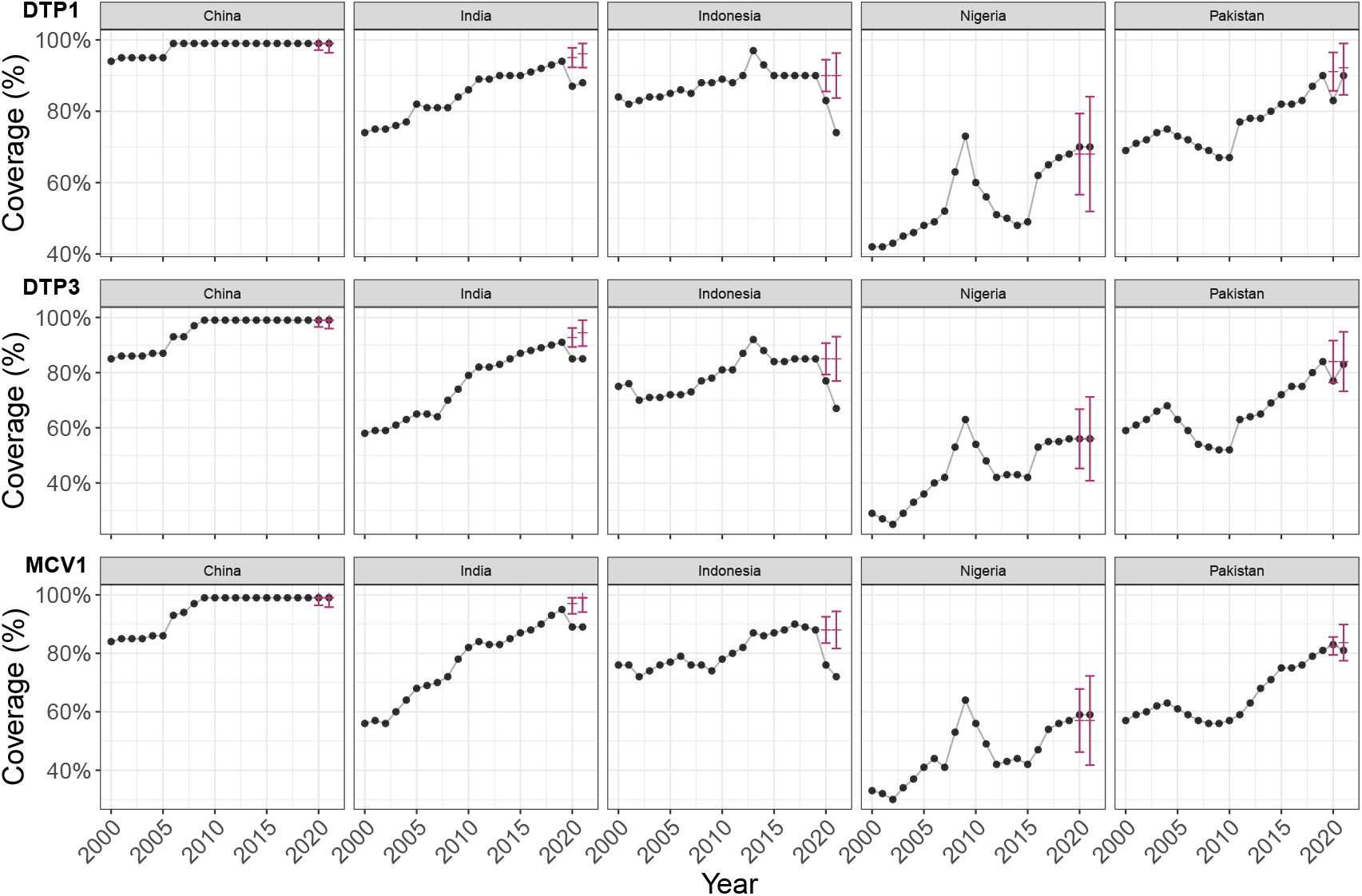
Expected and reported 2020 and 2021 vaccine coverage for DTP1, DTP3, and MCV1. example of five countries with largest populations in completed datasets. These graphs show WUENIC-reported coverage data (black dots) from 2000 to 2021 inclusive, and the corresponding ARIMA predictions and the associated 95% CIs (red bars) for 2020 and 2021.

The exact magnitude of coverage change remained hard to assess for two-thirds of countries due to uncertainties in model predictions (**Figure 2**) – however global trends remained clearly apparent. In 2021, eight more countries showed statistical evidence (p < 0.05) of changes in coverage compared to 2020 (for DTP3: n = 39 in 2021 and n = 31 in 2020. In 2020 all 39 countries had coverage declines below CIs and in 2021 30/31 countries had coverage declines below CIs, and 1 – Namibia - increased coverage above expected CIs). Conducting a *t-test* on within-country differences in coverage between years further confirmed that coverage declines were greater in 2021 than in 2020 (df = 160, t = -3.31, p = 0.0011).

**Figure 2.**
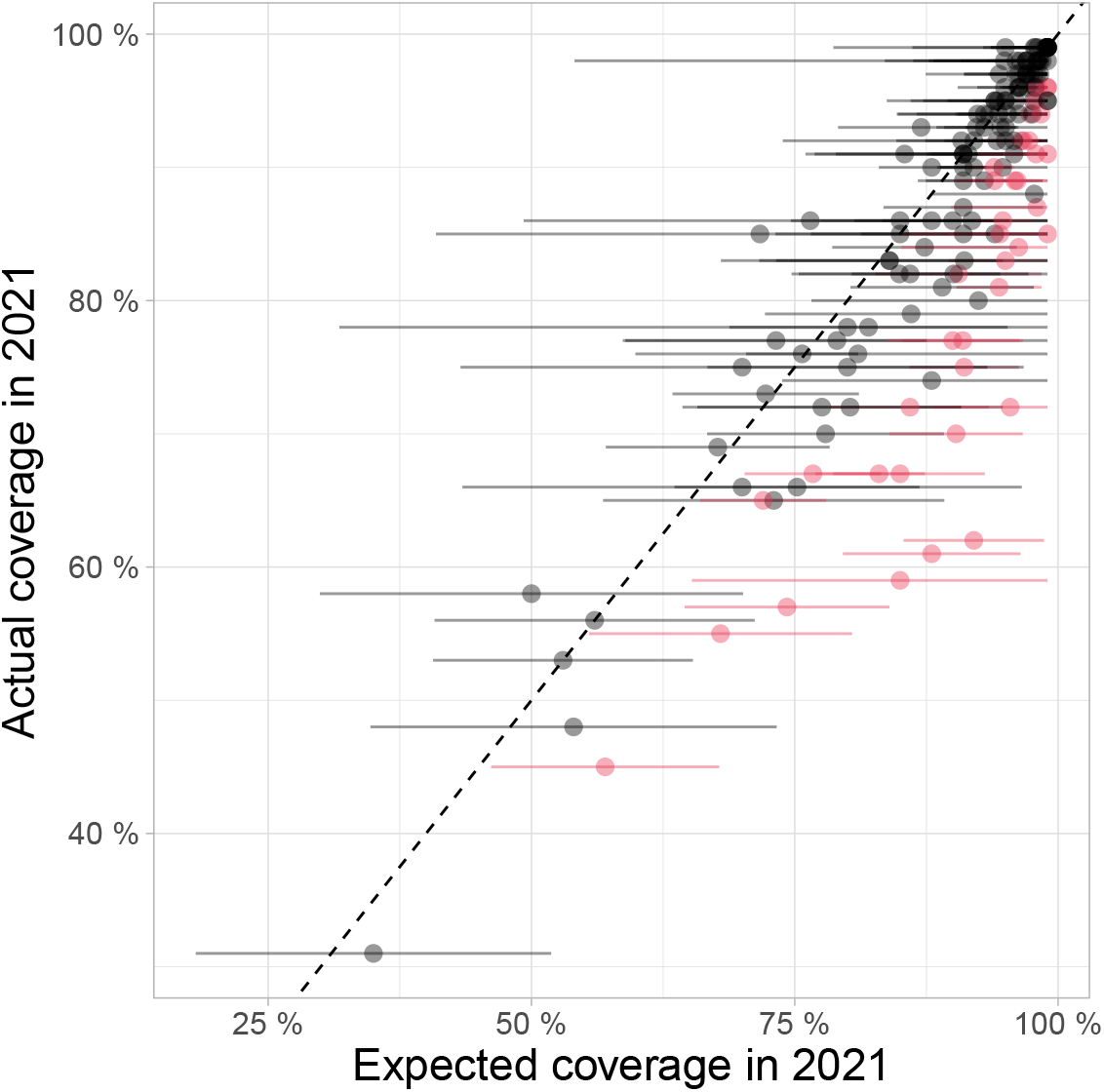
Comparison between 2021 WUENIC-reported DTP3 coverage and expectations derived from historical trends. This scatterplot shows country coverage (WUENIC-reported actuals and ARIMA-predicted expectations) as dots. Lines around individual points illustrate the 95% CIs of ARIMA predictions. Countries showing significant departure from expected values, *i*.*e*., for which actual coverage is outside the 95% CI of predictions, are indicated in red; countries without such significant departure from expected results are shown in black.

**Figure 3.**
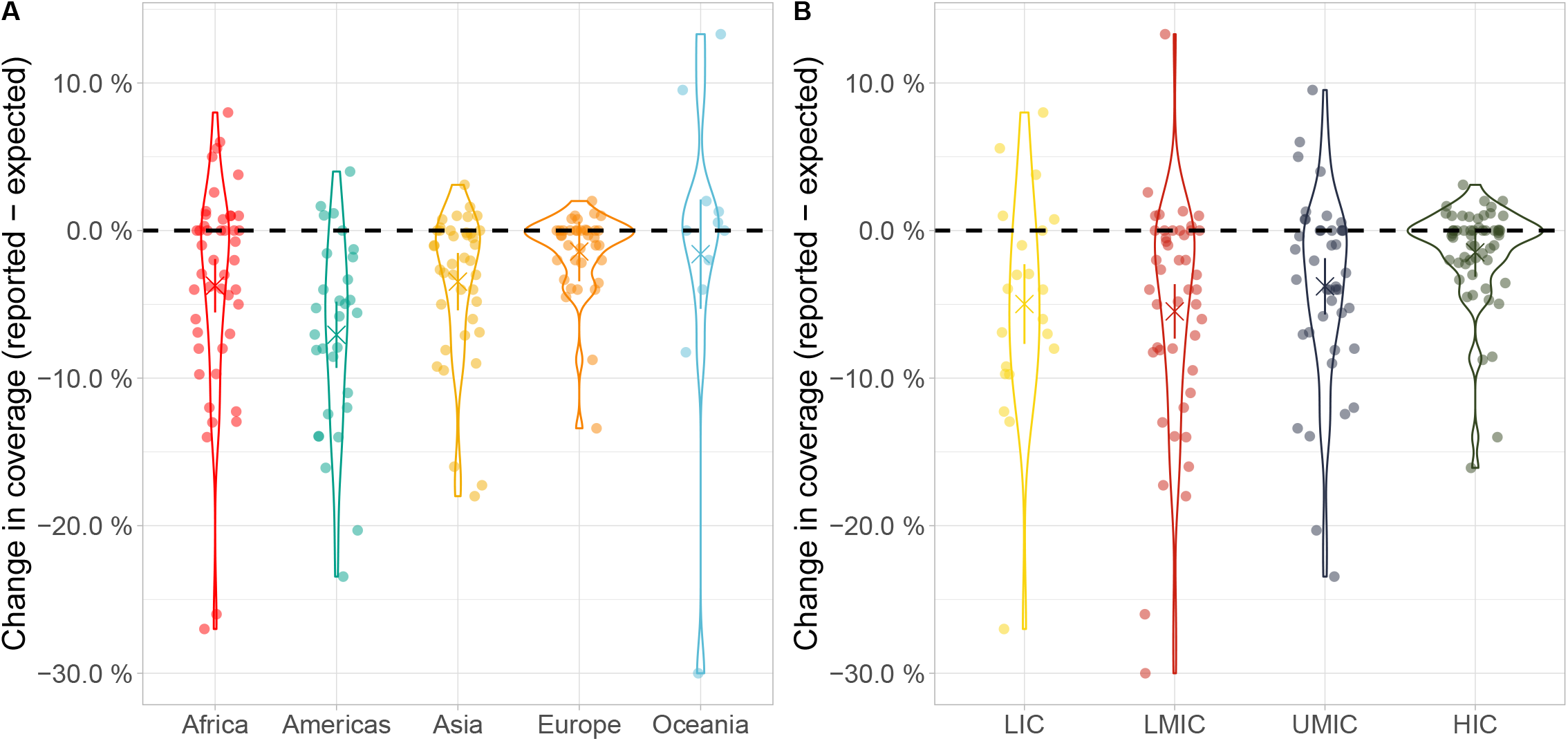
Differences between expected and reported DTP3 vaccine coverage in 2021 across (A) UN regions and (B) income groups. Points represent individual countries, grouped, and coloured according to (A) UN region classification and (B) World Bank income groups. Country coordinates on the X-axis were jittered for visibility. Values on the y-axis are indicated as absolute differences between reported and expected vaccine coverage, in percentages. Violin plots show the density of the data within each group: wider lines indicate more datapoints. The black dashed horizontal lines indicate no change in coverage. LIC: Low-income Country. LMIC: Lower-middle-income Country. UMIC: Upper-middle-income Country. HIC: High-income Country.

Our analysis indicates an average global decline in DTP3 coverage of 3·6% (95%_CI_: [2·6%; 4·6%]), from an expected 90·1% to a reported 86·5% across 164 reporting countries. Global average coverage in 2021 for this set of countries was last reported in 2005, continuing to validate our previously reported results that the pandemic is associated with a 16-year setback in performance of immunisation programmes globally. Similar trends were observed for DTP1 (global decline of 2·8%; 95%_CI_: [2·0%; 3·6%] from an expected 93·7% to a reported 90·9%), and for MCV1 (global decline of 3·8%; 95%_CI_: [4·8%; 2·7%]) in 2021. Full results by country and vaccine dose can be seen in the Appendix (Sections S5, S6, and Supplementary Tables). Sensitivity analyses also showed similar results (Appendix, Section S7).

Reported 2021 coverage represents a continued 0.8% decline from 2020 reported levels (DTP3: 87·3% in 2020 and 86·5% in 2021). Given the inclusion of historical updates to prior year coverage estimates for some countries, we re-evaluated our previously published analysis of 2020 coverage declines (we also updated inclusion and exclusion criteria, for consistency). Our re-analysis aligned with our previously reported results – DTP3 global decline of coverage of 2·7% (95%_CI_: [1·9%; 3·5%]), from an expected 90·0% to a reported 87·3%. Reproduced results are in the Appendix (Section S8).

At a regional level, there was evidence that coverage declined compared to previous trends in Africa, the Americas and Asia and that coverage in the Americas declined furthest (Americas: 6·7% decline, 95%_CI_: [4·8%; 9·3%]). Similar trends were seen across all vaccines modelled.

All income groups except for high-income countries exhibited statistical evidence of declines in coverage at the 95% confidence level in 2021 for DTP1, DTP3, and MCV1, with the great divergence in expected versus reported coverage consistently seen in lower-middle income countries (DTP3 LMICs; 5·5%, 95%_CI_: [3·6%; 7·3%]).

Heterogeneities due to one variable (regions or income groups) remained after accounting for the effect of the other one (regional after accounting for differences in income group: for DTP3, ANOVA – *F* = 5.22, df = 156, *p* < 5·63 × 10^−4^; income group after accounting for differences in regions: ANOVA – *F* = 5.75, df = 156, *p* < 9·33 × 10^−4^). Evidence of heterogeneities remaining after accounting for order effects were also seen for DTP1 (*p* = 0.018 for region after income, and *p* = *2.68* × 10^−4^ for income after region) and MCV1 (*p* = 0.003 for both ANOVAs).

### Additional missed immunisations and Zero Dose children in 2021

Zero Dose children are estimated using DTP1 data. These results on 168 countries for DTP1 represent 93% of the global surviving infant population in 2021. Based on point estimates of the total infant populations across these countries, there were 1.61 million (M) Zero Dose children in 2021, of which 504.9 k (31.4%) were associated with unexpected coverage declines in 2021. This point estimate is higher than the estimated additional 1.51 M ZD children in 2020, of which 358.3 k (23.8%) were associated with coverage declines in 2020 vs. prior trends.

**Table 1** details the number of missed immunisations for the 10 countries with the absolute number of additional missed immunisations compared to expectations for 2020 and 2021 combined. Of note, these countries are all in Asia, Africa, or the Americas. This list of countries is very similar to the list of countries with largest increases in missed immunisations in 2021 alone.

**Table 1.**
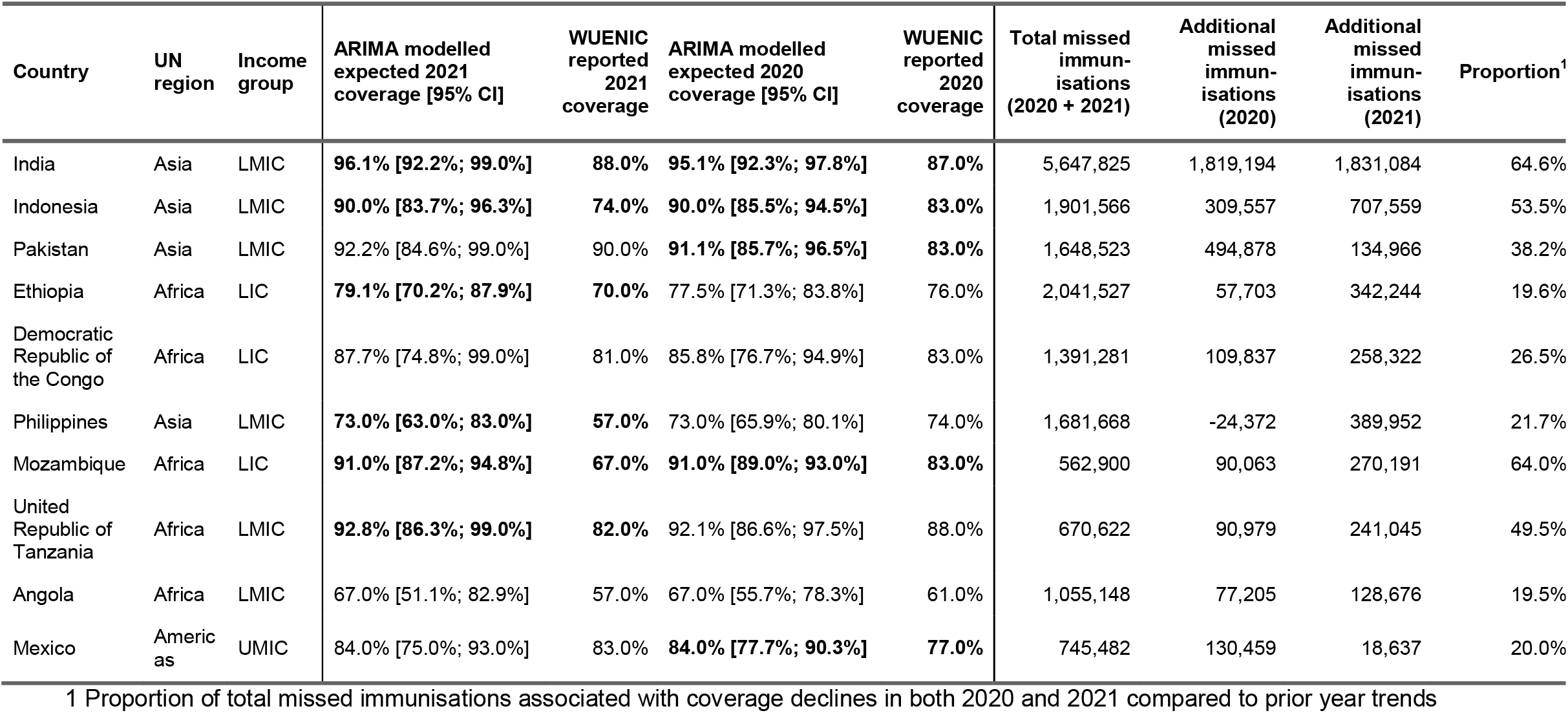
Estimated DTP3 coverage declines and missed immunisations for 10 countries with most additional missed immunisations total during 2020 and 2021. Bold rows indicate that WUENIC-reported coverage for each year is outside of the forecast 95% CIs.

| Country | UN region | Income group | ARIMA modelled expected 2021 coverage [95% CI] | WUENIC reported 2021 coverage | ARIMA modelled expected 2020 coverage [95% CI] | WUENIC reported 2020 coverage | Total missed immunisations (2020 + 2021) | Additional missed immunisations (2020) | Additional missed immunisations (2021) | Proportion<sup>1</sup> |
| --- | --- | --- | --- | --- | --- | --- | --- | --- | --- | --- |
| India | Asia | LMIC | **96.1% [92.2%; 99.0%]** | **88.0%** | **95.1% [92.3%; 97.8%]** | **87.0%** | 5,647,825 | 1,819,194 | 1,831,084 | 64.6% |
| Indonesia | Asia | LMIC | **90.0% [83.7%; 96.3%]** | **74.0%** | **90.0% [85.5%; 94.5%]** | **83.0%** | 1,901,566 | 309,557 | 707,559 | 53.5% |
| Pakistan | Asia | LMIC | 92.2% [84.6%; 99.0%] | 90.0% | **91.1% [85.7%; 96.5%]** | **83.0%** | 1,648,523 | 494,878 | 134,966 | 38.2% |
| Ethiopia | Africa | LIC | **79.1% [70.2%; 87.9%]** | **70.0%** | 77.5% [71.3%; 83.8%] | 76.0% | 2,041,527 | 57,703 | 342,244 | 19.6% |
| Democratic Republic of the Congo | Africa | LIC | 87.7% [74.8%; 99.0%] | 81.0% | 85.8% [76.7%; 94.9%] | 83.0% | 1,391,281 | 109,837 | 258,322 | 26.5% |
| Philippines | Asia | LMIC | **73.0% [63.0%; 83.0%]** | **57.0%** | 73.0% [65.9%; 80.1%] | 74.0% | 1,681,668 | -24,372 | 389,952 | 21.7% |
| Mozambique | Africa | LIC | **91.0% [87.2%; 94.8%]** | **67.0%** | **91.0% [89.0%; 93.0%]** | **83.0%** | 562,900 | 90,063 | 270,191 | 64.0% |
| United Republic of Tanzania | Africa | LMIC | **92.8% [86.3%; 99.0%]** | **82.0%** | 92.1% [86.6%; 97.5%] | 88.0% | 670,622 | 90,979 | 241,045 | 49.5% |
| Angola | Africa | LMIC | 67.0% [51.1%; 82.9%] | 57.0% | 67.0% [55.7%; 78.3%] | 61.0% | 1,055,148 | 77,205 | 128,676 | 19.5% |
| Mexico | Americas | UMIC | 84.0% [75.0%; 93.0%] | 83.0% | **84.0% [77.7%; 90.3%]** | **77.0%** | 745,482 | 130,459 | 18,637 | 20.0% |

1 Proportion of total missed immunisations associated with coverage declines in both 2020 and 2021 compared to prior year trends

### Year-on-year trends in point estimates of coverage

Absolute reported coverage indicates the proportion of children immunised in a given year per country, and can be compared to previous coverage levels to understand if more or less children are being reached each year. In terms of absolute reported coverage from 2019 to 2021, for DTP3, 40 countries (24.4%) reported continued coverage declines for both consecutive years whereas just six countries (3.7%) reported sustained coverage improvements year-on-year. The majority of countries either exhibited coverage stagnation (39 countries, 23.8%) or a decline follow-up by stagnation (or stagnation followed by a decline) - 24 countries (14.6%). The remaining 48 countries (29.3%) had coverage oscillate (e.g., lower then higher, or higher then lower) over years. **Table 2** for a full breakdown for DTP3; similar trends were seen for DTP1 and MCV1.

**Table 2.**
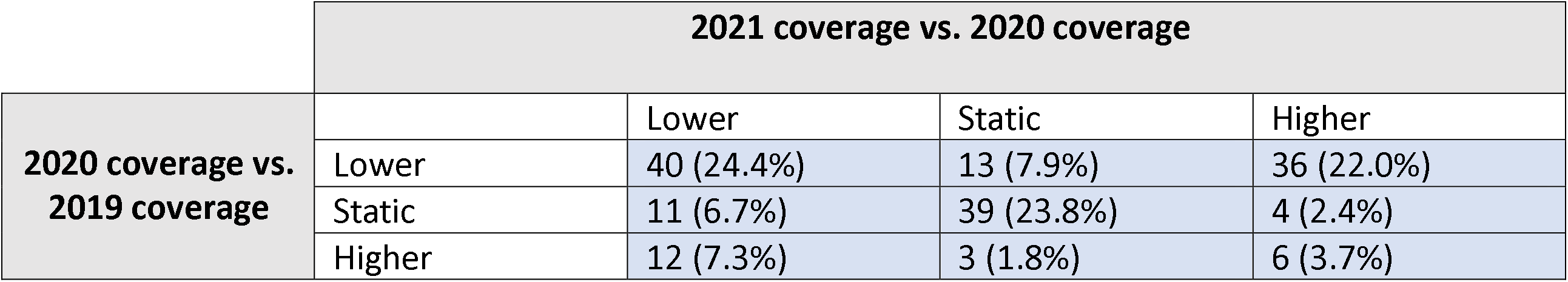
Tabulation of number of countries by absolute year-on-year coverage trends for DTP3. Percentages show percent of all 164 included countries.

At a country level the year-on-year reported changes in coverage can be seen in **Figure 4,** with Panel A providing colour-coding by region and panel B by income group. Only six countries are in the top right segment (consistent year-on-year coverage improvement for DTP3 during the first two years of the pandemic) and all other countries reported some declines and/or static coverage levels during the pandemic.

**Figure 4.**
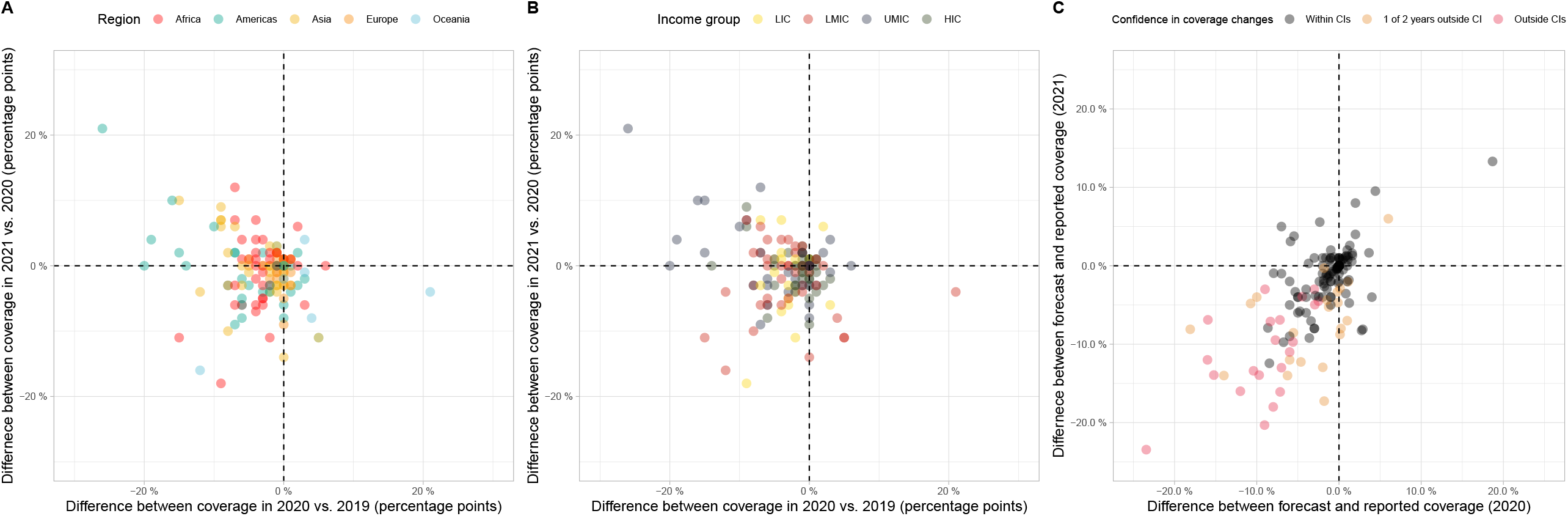
Year-on-year DTP3 trends: Comparison between 2021 WUENIC-reported DTP3 coverage and expectations derived from historical trends. Scatterplots A and B show the difference between 2020 and 2019 WUENIC-reported coverage on the x-axis, and 2021 and 2020 WUENIC-reported coverage on the y-axis, categories by region (A) and income group (B) respectively. Countries in the top-right corner increased coverage on both sequential years, whilst countries in the bottom-left countries reported year-on-year declines. Scatterplot C shows the difference between forecasted and reported coverage in 2020 on the x-axis and in 2021 on the y-axis. Countries in the top-right corner reported higher coverage than expected for both years, whilst countries in the bottom-left corner reported lower coverage than expected for both years. Colour-coding indicates whether reported coverage was within CIs of modelled projections or not. All figures show percentage point differences.

Year-on-year trends compared to forecasts

Comparing reported coverage to forecasted levels provides an indication of whether, and to what extent, coverage deviated from country-specific previous trends during each year of the pandemic. For the majority of countries (118, 72.0%) reported coverage fell within 95% CI forecasts for both 2020 and 2021. No countries had higher coverage in 2021 than forecasted confidence intervals, however 24 countries (14.6%) had lower coverage than expected in both years. A Fisher’s Exact test confirmed that trends observed in 2020 and in 2021 were correlated (DTP3: *p* < 2 × 10^−13^), largely due to an access of countries exhibiting declines in 2020 which continued in 2021 (**Table 3**)

**Table 3.** Tabulation of number of countries comparing reported vs. expected coverage in 2020 and 2021 for DTP3. Percentages show percent of all countries.

|  |  | 2021 reported coverage vs. expectations |  |  |
| --- | --- | --- | --- | --- |
| 2020 reported coverage vs. expectations |  | Lower | Within CIs | Higher |
|  | Lower | 24 (14.6%) | 6 (3.7%) | 0 (0%) |
|  | Within CIs | 15 (9.1%) | 118 (72.0%) | 0 (0%) |
|  | Higher | 0 (0%) | 1 (0.6%) | 0 (0%) |

## Discussion

### Global routine immunisation continues decline

Our results suggest that the global decline in RI coverage observed in 2020 has continued in 2021. In contrast with previous indications in the latter half of 2020 that services and coverage was returning to pre-pandemic levels [22], we found virtually no indication of rebounds in RI coverage, and many instances of countries where decline continued further in 2021 (n = 40, 24.4% of countries), where declines occurred following stagnation in the first year of the pandemic (*n* = 11, 6.7% of countries), or coverage continues to stagnate at 2019 levels (*n* = 39, 23.8% of countries). Immunisation performance has not reverted to pre-pandemic levels during the continuation of the pandemic and has instead declined further from the previously achieved improvement trajectory. Global WUENIC-reported coverage across the 164 included countries for DTP3 declined continuously from 89.8% in 2019 to 87.3% in 2020 to 86.5% in 2021. This compares to an expected 90·1% coverage in 2021 (*i*.*e*., decline of 3·6%, 95%_CI_: [2·6%; 4·6%]). See **Figure 5** for an overview of DTP1, DTP3, and MCV1 coverage actuals and expectations from 2000 to 2021.

**Figure 5.**
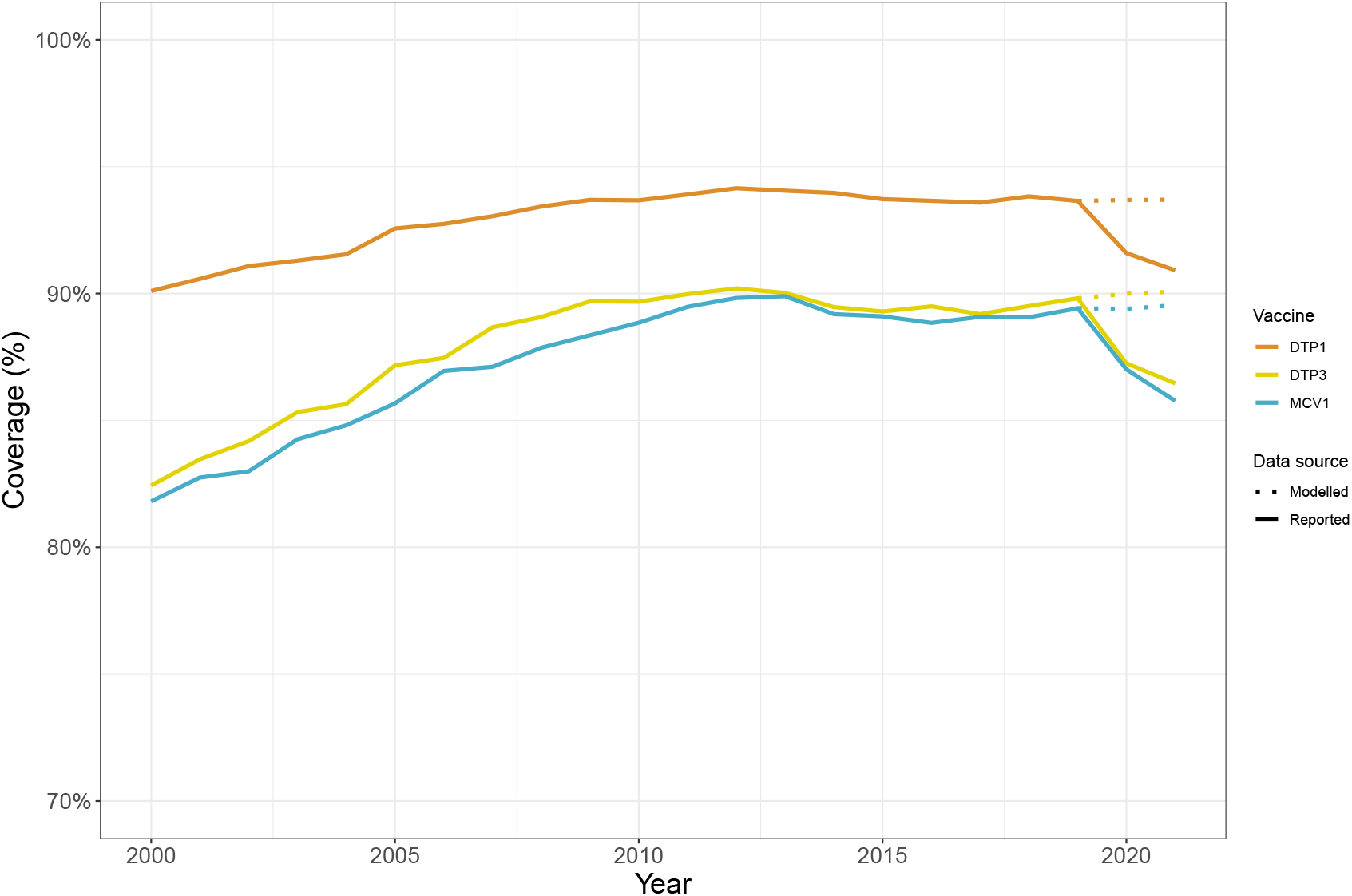
Global DTP1, DTP3, and MCV1 coverage. Line graph shows DTP1 coverage in red, DTP3 coverage in green, and MCV1 coverage in blue from 2000-2021 inclusive. Filled lines show reported WUENIC coverage and dotted lines from 2019-2021 show expected trends based on ARIMA modelling of prior coverage.

Between 2019 and 2021, two-thirds of countries exhibited either continuous declines year-on-year, or declines followed by stagnation (or vice versa) during the pandemic. These findings confirm qualitative reports from the World Health Organisation (WHO) that immunisation services have been continuously interrupted throughout the pandemic (56% of countries reported disruptions in Q3 2020, 42% in Q1 2021, and 53% in Q4 2021 [23]).

The trend in declines in RI coverage during the pandemic period is largest in the Americas, Africa, and Asia; and for low- and middle-income countries (with region and income group both having independent explanatory power). This trend is also consistent across vaccines, delivered at different schedules (DTP1 typically delivered at 6-weeks, DTP3 at 14-weeks, and MCV1 at 9-months old).

Our results cover over 90% of the surviving infant population in 2021 – over 119 M infants. However, our results are limited to annual trends reported at a national level. Publicly available data on globally recognised estimates of coverage at a monthly level would enable more granular time series modelling on the impact of the pandemic on RI, ideally with shorter time lags between events and reporting.

### Missed immunisations

The compounded impact of two years with RI coverage at lower levels (or coverage declines then stagnation or vice versa) in many countries will place an increasing toll on RI vaccine requirements and increase complexity of service delivery. In 2023, there will be an increasing number of unvaccinated one- and two-year-olds in many countries. Vaccine programmes typically plan vaccine supply requirements and procurement based on surviving infant populations. This approach may need to evolve to actively include quantification of missed immunisation in older populations to ensure sufficient vaccines for catch-up through RI touchpoints. Furthermore, reaching children over one-years old is not “typical” (in most countries) for RI, and infants of this age do not necessarily routinely engage with the health system at appropriate times and places to be reached through ad-hoc catch-up. Resultingly, catch-up campaigns – targeted at sub-populations where coverage has declined the most – may be required. More data and research at the sub-national level is required in order to determine the most appropriate catch-up routes and supplementary immunisation activities (SIAs). WUENIC data is published at national level, therefore alternative coverage data sources, e.g., administration data (which has some known caveats – see Appendix Section S1) or other sub-national level surveys would be required.

### Zero Dose

Notably, the number of Zero Dose children also continued to grow: DTP1 coverage across the modelled countries fell from a reported 91.6% to 90.9% between 2020 and 2021, compared to an expected 93·7% coverage in 2021 (*i*.*e*., decline of 2·8%, 95%_CI_: [2·0%; 3·6%]) - see **Figure 5** for visual depiction. This translates to an additional 3.4 million Zero Dose children on top of an expected 11.0 million (30.9% increase). These figures for both 2020 and 2021 are updated using the most recent UN WPP estimates, based on the medium variant surviving infant population estimates. Population estimates, particularly in fragile states where a national census may not have been conducted for years or decades, are known to sometimes have wide confidence intervals (see UN WPP 2022 projections) - therefore the number of Zero Dose children could be larger still.

The decline in DTP1 coverage was 77% of the size of the decline in DTP3 coverage (2.8% vs. 3.6%). This suggests that of the additional 4.3 M infants not receiving DTP3 globally in 2021 associated with pandemic declines, 3.3 M of these were additional Zero Dose children versus expectations, and 1 M resulted from increased drop out between vaccine doses. On the one hand, this is particularly concerning given the tight linkage between Zero Dose children and adverse health outcomes (e.g., disease outbreaks and vaccine preventable deaths). However, recognition that declines in coverage appear to be primarily driven by increases in ZD, rather than greater dropout between vaccine doses, provide insights on where to target limited resources for catch-up on health system strengthening. This places heightened importance on the increasing focus on channeling resources to identify and reach Zero Dose children.

Additionally, several individual countries with large birth cohorts reported coverage below the expected 95% CIs, indicating strong evidence in large increases in Zero Dose children during the continued pandemic. These countries include: India, Indonesia, Philippines and Mozambique (see **Table 1** above) - and these could be important focus countries for health system strengthening support.

### Vaccine preventable disease outbreaks

The large decline in Measles coverage (MCV1 global decline of 3·8%; 95%_CI_: [4·8%; 2·7%]) is particularly concerning given the high sustained coverage (95%) required to prevent measles outbreaks [24]. Global disease surveillance suggests that this – along with other factors including delays to measles campaign implementation, potentially also pandemic related – are translating into an increase in worldwide measles outbreaks [25]. Recent modelling has modelled the link between declines in immunisation coverage and vaccine-preventable disease outbreaks based on scenarios of uniform declines in coverage [26] - such methods could be enhanced to build on our country-specific modelled declines in coverage to more precisely estimate potential increases in vaccine-preventable diseases. This could help proactively target campaigns where outbreaks would be most likely to occur, if real time coverage declines are identified.

### Factors for pandemic resilience

Whilst income group and regional factors (independently) explain some of the global declines in coverage (adjusted R^2^ = 0.14 for the linear model including income group and region), further understanding of explanatory factors is required to guide recovery and build resilience globally for future pandemics. Understanding the impact of Non-Pharmaceutical Interventions (NPIs) and COVID-19 rollout on health service access and delivery may be valuable starting points. Countries deployed a wide variety of NPIs over time, which influenced mobility and societal behaviours [27], and influenced immunisation services [28]. These interventions have been captured and codified in global datasets, such as the COVID-19 Government Response Tracker [29]. Exploring the relationship between NPI implementation and immunisation system performance over the last two years may indicate policy areas with beneficial and deleterious effects on health systems. Additionally, approximately 50% of countries surveyed by the WHO reported direct trade-offs between COVID-19 vaccination programme rollout and RI service delivery for infants and school-aged children [23]. Further quantification of the trade-off between continued COVID-19 vaccination efforts and RI, in terms of morbidity and mortality implications, could be valuable in guiding immunisation strategies at the country- and global-level in the coming years (as efforts to integrate COVID-19 in immunisation programmes continue).

Finally, a select few examples of countries achieving coverage improvements year-on-year and/or reported coverage higher than forecasts in 2020, despite the backdrop of the pandemic. These countries may provide case studies to explore for translational learnings. For example, Chad’s reported DTP3 coverage increased from 50% in 2019 to 52% in 2020 and 58% in 2021 (although it is noted that coverage was within 95% forecasted CIs for both years); and Namibia’s 2020 reported DTP3 coverage of 93% was higher than forecasted (point estimate: 87% (95%_CI_: [81·4%; 92·6%]).

## Conclusion

This research builds on our previously published, transparent, and replicable approach for estimating gaps in RI coverage across countries, providing an objective measure for missed immunisations and coverage disruptions in 2021. It is clear that immunisation services continued to be disrupted in many low- and middle-income countries, particularly in the Americas, Africa, and Asia, and we hope this work can inform future research to identify effective interventions to facilitate rebounds in coverage to previous levels and catch-up of growing populations of under- and un-immunised children.

## Data Availability

All data produced are available online on GitHub - https://github.com/bevans249/modelling_covid_impact_RI

## Acknowledgements

TJ acknowledges funding from the MRC Centre for Global Infectious Disease Analysis (reference MR/R015600/1), jointly funded by the UK Medical Research Council (MRC) and the UK Foreign, Commonwealth & Development Office (FCDO), under the MRC/FCDO Concordat agreement and is also part of the EDCTP2 programme supported by the European Union. OK was funded by the Swiss National Science Foundation (grant number PP00P3_202660). These funders had no role in the design and conduct of this study; collection, management, analysis, and interpretation of the data; preparation, review, or approval of the manuscript; and decision to submit the manuscript for publication.

BE is a PhD student at the Institute of Global Health, Faculty of Medicine, University of Geneva and received no funding.

