## Supplementary text for "Time series analysis of routine immunisation coverage during the COVID-19 pandemic in 2021 shows continued global decline and increases in Zero Dose children"

**Supplementary Material: Additional data, methodological information, and complementary results**

**Captions:**

- Supplementary material (Word file): This supplementary text provides further details on: selection of datasets, methodologies used for modelling, countries/ datapoints removed from analyses. It also provides full outputs across all countries, and links to the GitHub repository for original datasets and R code.
- Supplementary Tables (Excel file): This table includes results for all countries for DTP1, DTP3, and MCV1 modelling for 2020 and 2021, in an accessible Excel format. There is one tab for each vaccine dose, and details in the Word document on how to interpret.

**Contents:**

Section S1: Vaccination coverage data selection

Section S2: Further datasets for categorising countries

Section S3: Using ARIMA to model expected vaccination coverage

Section S4: Countries removed from analyses

Section S5: Full country-level results for 2020 and 2021

Section S6: Summary income group and region results for 2021

Section S7: Sensitivity analyses for 2021

Section S8: Reproduced global 2020 results

Section S9: Availability and reproducibility

**Section S1: Vaccination coverage data selection**

Vaccine coverage is typically estimated through (a) aggregating (raw) administration data, or (b) conducting surveys. Expert opinion and/ or statistical methods may be layered on to produce final estimates. Administration data is the most timely and periodic, but risks numerator (e.g., under- or over-estimation dependent on health system capacity, reporting incentives, and linkage to private sector systems) and denominator (e.g., out-of-date censuses used for population quantification) biases [1]. Surveys, typically household or parental, avoid such biases but are expensive, time-consuming, infrequent, and may encounter recall bias, i.e., parents or guardians may mis-remember or confuse vaccination statuses when reporting due to complex immunisation schedules and potentially long periods between delivery and surveys [2]. The Institute for Health Metrics and Evaluation (IHME) produces coverage estimates by applying spatiotemporal statistical methods to household survey microdata (if available) and estimates of country-reported coverage data [3]. IHME usefully publish confidence ranges, unlike other sources, but methods are not fully reproducible nor used routinely by global immunisation stakeholders to assess immunisation performance. WUENIC estimates are published annually through computational logic rule-based approaches that use inputs from country-reported administrative and survey data, adjusted based on expert assessment and country consultations [4], [5]. WUENIC estimates are transparent, replicable, and routinely used by stakeholders to inform policy, financial, and programmatic decisions.

WUENIC data was selected due to its transparency, replicability, utility by key organisations and donors working on or investing in immunisation globally (e.g., UNICEF, WHO and Gavi), and public availability. From this data, three RIs – DTP1, DTP3 and MCV1 – were selected since they act as key immunisation indicators.

**Section S2: Further datasets for categorising countries**

Three additional sources were used to assemble country demographic information – population data from the United Nations World Population Prospects (UNWPP [6]), World Bank (WB) income group classification [7], and United Nations (UN) regional classifications (using the “countrycode” package in R, [8]).

**Section S3: Using ARIMA to model expected vaccination coverage**

WUENIC does not publish future-looking coverage forecasts. Expected coverage for 2020 and 2021, by country in the absence of COVID-19 was modelled by fitting AutoRegressive Integrated Moving Average (ARIMA) models to 20-years of annual WUENIC coverage data (2000-2019) using the package “*forecast*”) in R [9] and projecting models forward for two years. No projections were modelled for countries missing any coverage values from 2000 to 2019.

For each time series the appropriate ARIMA model based on historic coverage trends was automatically selected using the function “*auto.arima*” in the “*forecast”* package [9]. ARIMA models are characterised by three order terms – *p*, *d*, and *q* – representing the order of the Auto Regressive (AR) term, the number of differencing steps required to make the time series stationary (Integrated, I, term), and the order of the Moving Average (MA) term respectively. Since WUENIC data has annual periodicity, there are no seasonal patterns, and seasonal ARIMA models were not required. Automatic ARIMA modelling fits the appropriate *p*, *d*, and *q* order terms based on:

- Conducting Kwiatkowski-Phillips-Schmidt-Shin tests, with the null hypothesis that the time series is stationary around a deterministic trend against the alternative of a unit root to validate whether the time series is stationary [10]. Where not stationary, each time series is differenced and then re-validated to test if stationary to determine the integrative order, *d*.
- Stepwise algorithm to traverse the model space to select the model with the smallest Akaike Information Criterion (AIC) [11]. AIC scores evaluate how well a model fits the data it was generated from, and lower AIC scores (for the same dataset) indicate better model fit. AIC minimisation identifies the *p* and *q* terms.

The mean ARIMA-modelled value was selected per country and vaccine as expected 2020 coverage. WUENIC cap coverage at 99% [4], [5]. To avoid falsely calculating small declines in coverage for countries where ARIMA-predicted 2020 coverage was higher than 99%, all expected coverage estimates were capped at the WUENIC maximum.

**Section S4: Countries removed from analyses**

Before further analysis, coverage time series including expected coverage were investigated to identify unreliable estimates as described in the manuscript.

The following countries were removed from all analyses:

(1) Countries lacking recent WUENIC coverage updates: Cambodia, the Central African Republic, Haiti, Guinea, Lesotho, Somalia, and South Sudan
(2) Countries with major geopolitical events: Myanmar, Democratic People’s Republic of Korea
(3) Poor model fit:
    - **DTP1**: Barbados (Plurinational State of), Chile, Guatemala, Iraq, Jordan, Kiribati, Lesotho, Libya, Samoa, Serbia, Solomon Islands, South Africa, Uzbekistan, Viet Nam
    - **DTP3**: Argentina, Belarus, Brazil, El Salvador, Iraq, Jordan, Kiribati, Lesotho, Libya, Lithuania, Malawi, Malta, Mauritania, Palau, Serbia, Solomon Islands, Turkmenistan, Venezuela (Bolivian Republic of), Yemen
    - **MCV1**: Andorra, Angola, Antigua and Barbados, Argentina, Bahamas, Bolivia (Plurinational State of), Cote d’Ivoire, Gambia, Honduras, Jordan, Kiribati, Latvia, Madagascar, Malaysia, Mexico, Palau, Philippines, Samoa, Serbia, Suriname, Trinidad and Tobago, Venezuela (Bolivian Republic of)

Coverage trends from 2000-2021 and modelled ARIMA expected coverage for 2020 and 2021, and associated confidence intervals, for each removed country can be seen in Figure S4.1 for DTP1, S4.2 for DTP3, and S4.3 for MCV1.

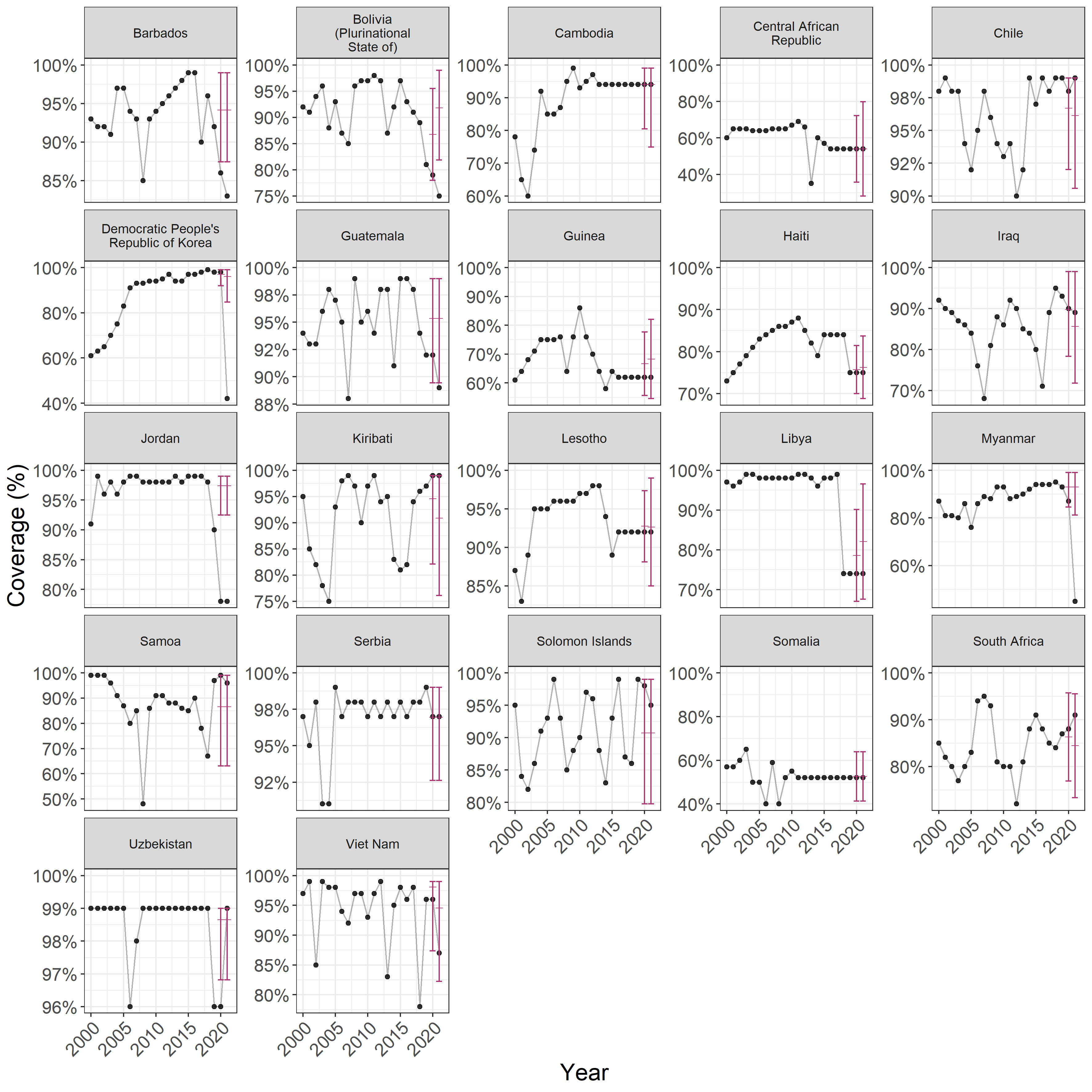
**Figure S4.1: Expected and reported 2020 and 2021 vaccine coverage for DTP1 for countries removed from aggregate analyses.** These graphs show WUENIC-reported coverage data (black dots) from 2000 to 2021 inclusive, and the corresponding ARIMA predictions and the associated 95% CIs (red bars) for 2020 and 2021.

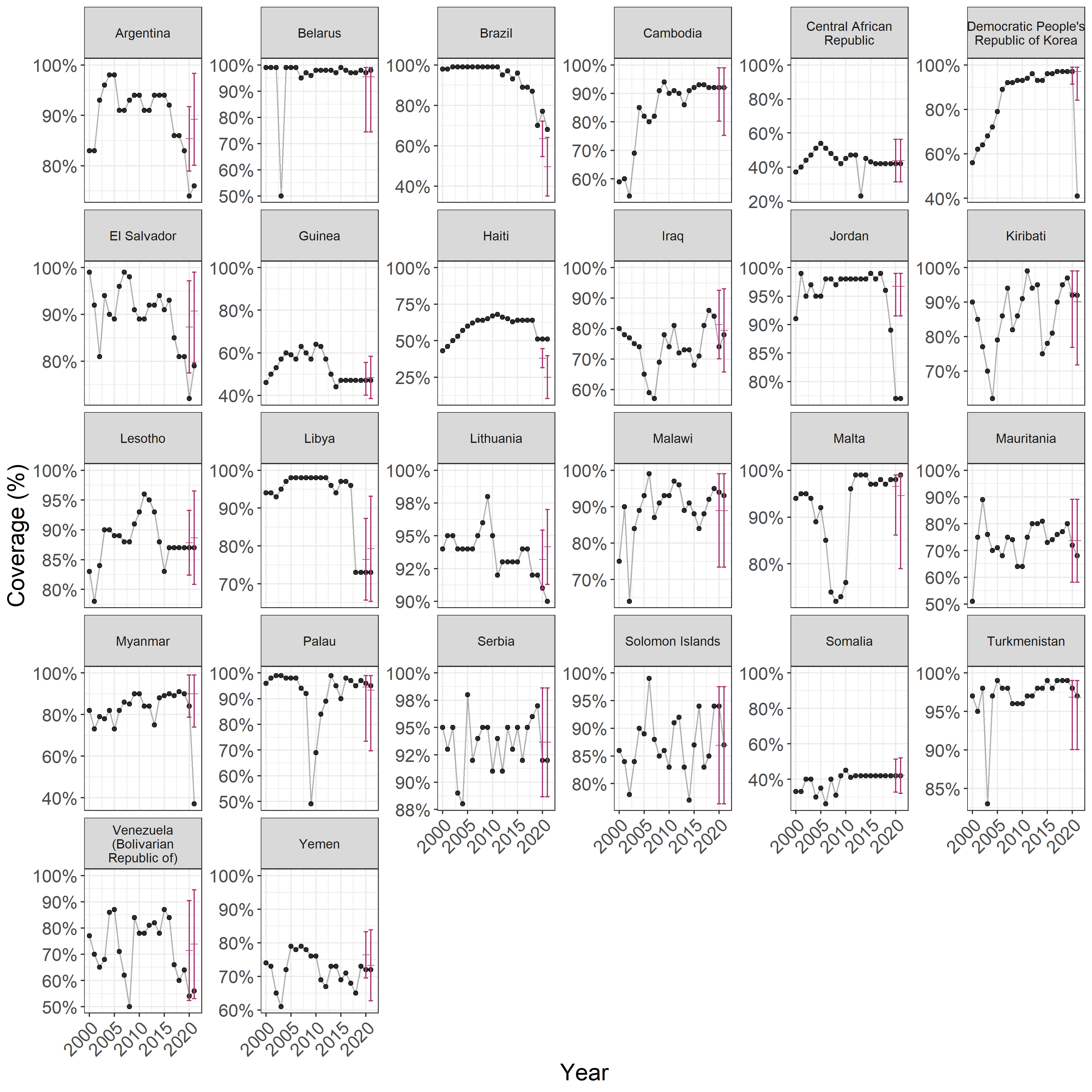

**Figure S4.2: Expected and reported 2020 and 2021 vaccine coverage for DTP3 for countries removed from aggregate analyses.** These graphs show WUENIC-reported coverage data (black dots) from 2000 to 2021 inclusive, and the corresponding ARIMA predictions and the associated 95% CIs (red bars) for 2020 and 2021.

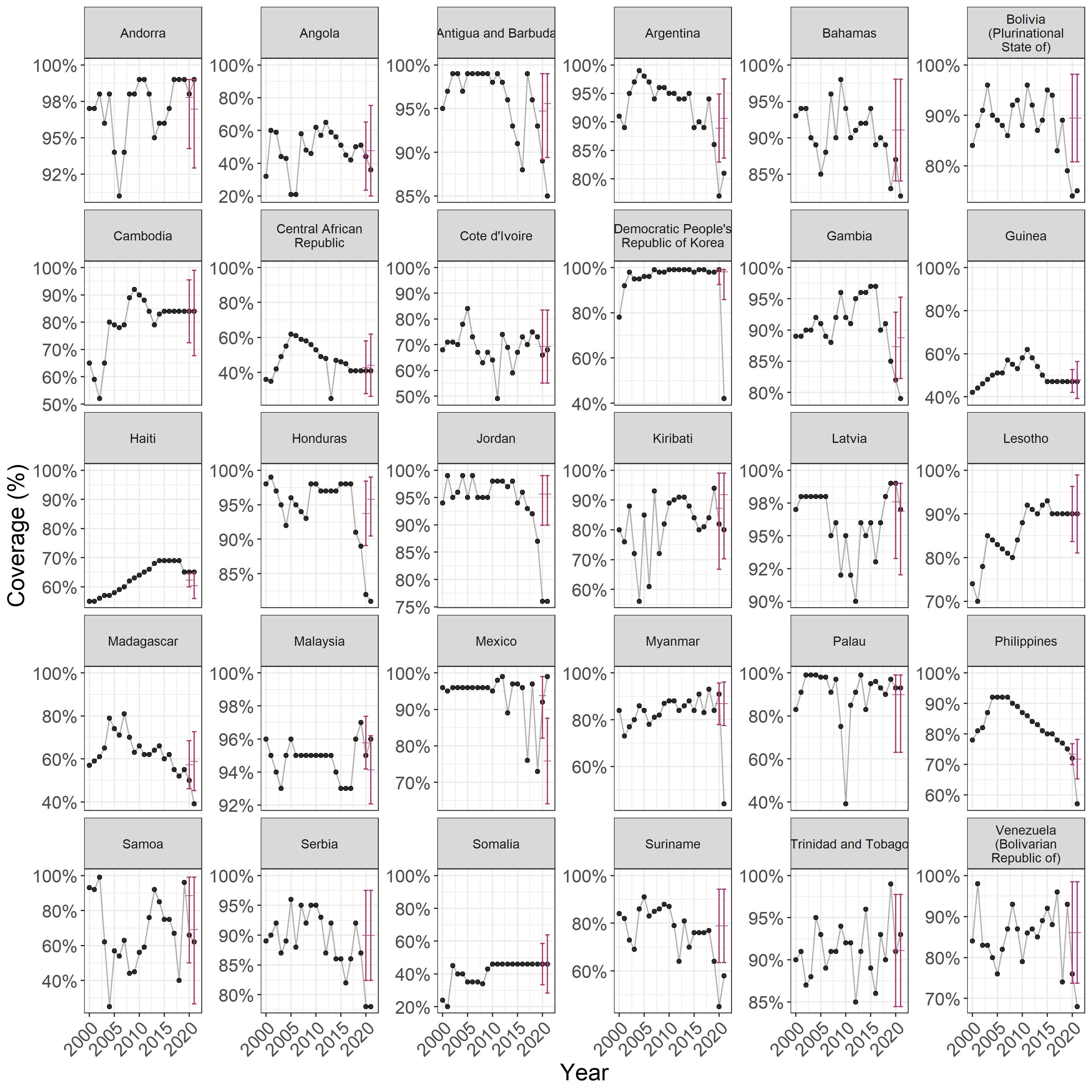

**Figure S4.3: Expected and reported 2020 and 2021 vaccine coverage for MCV1 for countries removed from aggregate analyses.** These graphs show WUENIC-reported coverage data (black dots) from 2000 to 2021 inclusive, and the corresponding ARIMA predictions and the associated 95% CIs (red bars) for 2020 and 2021.

**Section S5: Full country-level results for 2020 and 2021**

Full output results in a file format from which data can be efficiently extracted are available in **Supplementary Materials Table S1** for DTP1, and **Supplementary Materials Table S2** for DTP3, and **Supplementary Materials Tables S3** for MCV1. These tables include results for both 2020 and 2021:

- Country classification details: country, ISO code, UN region and income group
- ARIMA predictions: mean, low- and high- 95% confidence intervals, confidence interval width for 2020 and 2021
- WUENIC reported coverage: for 2020 and 2021
- Deltas (expected - reported coverage): mean, low- and high- 95% confidence intervals for 2020 and 2021
- 95% confident flag: binary flag whether WUENIC-reported 2020 coverage is outside the ARIMA-predicted 95% confidence intervals (CIs) each year. TRUE if within CIs, and FALSE if outside CIs.
- Surviving infant population: from UN WPP medium variant births minus infant mortality for each year
- Missed immunisation estimates: total missed immunisations (based on WUENIC-reported coverage), Expected (based on mean ARIMA prediction coverage) for each year
- Additional missed immunisation estimates: mean (based on mean coverage delta), low and high estimates (based on delta 95% confidence intervals) for each year
- Country removed from analysis flag: binary flag whether the country was removed from the aggregate (total, region, and income group) analyses. TRUE if removed, FALSE if included.

**Section S6: Summary global, income group and region results for 2021**

*S6(a) T-test of global impact – final results, 2021*

| **Vaccine** | **Sample size** | **WUENIC reported coverage** | **ARIMA expected coverage** | **Delta** | **95% CI (low)** | **95% CI (high)** | ***t* statistic** | ***p* value** |
| --- | --- | --- | --- | --- | --- | --- | --- | --- |
| **DTP1** | 168 | 90.9% | 93.7% | -2.8% | -3.6% | -2.0% | - 6.97 | 7.07E-11 |
| **DTP3** | 164 | 86.5% | 90.1% | -3.6% | -4.6% | -2.6% | - 7.20 | 2.11E-11 |
| **MCV1** | 160 | 85.8% | 89.5% | -3.8% | -4.8% | -2.7% | - 7.16 | 2.83E-11 |

*S6(b) T-test by region – final results, 2021*

(i) *DTP1*

| **Group** | **Sample size** | **WUENIC reported coverage** | **ARIMA expected coverage** | **Delta** | **95% CI (low)** | **95% CI (high)** | ***t* statistic** | ***p* value** |
| --- | --- | --- | --- | --- | --- | --- | --- | --- |
| **Africa** | 47 | 86.7% | 90.2% | -3.4% | -4.9% | -2.0% | - 4.68 | 5.94E-06 |
| **Americas** | 30 | 88.3% | 93.3% | -5.0% | -6.8% | -3.1% | - 5.38 | 2.52E-07 |
| **Asia** | 40 | 93.0% | 95.2% | -2.3% | -3.9% | -0.7% | - 2.88 | 0.005 |
| **Europe** | 40 | 96.1% | 97.0% | -0.9% | -2.5% | 0.7% | - 1.10 | 0.27 |
| **Oceania** | 11 | 89.8% | 92.5% | -2.7% | -5.7% | 0.3% | - 1.76 | 0.08 |

(ii) *DTP3*

| **Group** | **Sample size** | **WUENIC reported coverage** | **ARIMA expected coverage** | **Delta** | **95% CI (low)** | **95% CI (high)** | ***t* statistic** | ***p* value** |
| --- | --- | --- | --- | --- | --- | --- | --- | --- |
| **Africa** | 46 | 80.6% | 84.4% | -3.7% | -5.6% | -1.9% | - 4.08 | 7.20E-05 |
| **Americas** | 30 | 84.7% | 91.8% | -7.1% | -9.3% | -4.8% | - 6.22 | 4.21E-09 |
| **Asia** | 40 | 89.9% | 93.3% | -3.5% | -5.4% | -1.5% | - 3.52 | 5.64E-04 |
| **Europe** | 37 | 92.5% | 93.9% | -1.4% | -3.4% | 0.6% | - 1.39 | 0.17 |
| **Oceania** | 11 | 82.8% | 84.4% | -1.6% | -5.3% | 2.1% | - 0.85 | 0.40 |

(iii) *MCV1*

| **Group** | **Sample size** | **WUENIC reported coverage** | **ARIMA expected coverage** | **Delta** | **95% CI (low)** | **95% CI (high)** | ***t* statistic** | ***p* value** |
| --- | --- | --- | --- | --- | --- | --- | --- | --- |
| **Africa** | 45 | 78.3% | 82.3% | -3.9% | -5.8% | -2.0% | - 4.09 | 7.01E-05 |
| **Americas** | 25 | 85.2% | 93.0% | -7.8% | -10.4% | -5.3% | - 6.07 | 9.54E-09 |
| **Asia** | 41 | 90.9% | 93.3% | -2.5% | -4.5% | -0.5% | - 2.46 | 0.02 |
| **Europe** | 38 | 91.3% | 93.3% | -2.0% | -4.1% | 0.1% | - 1.90 | 0.06 |
| **Oceania** | 11 | 79.5% | 84.4% | -4.9% | -8.8% | -1.1% | - 2.53 | 0.01 |

*S6(c) T-test by income group – final results, 2021*

(i) *DTP1*

| **Group** | **Sample size** | **WUENIC reported coverage** | **ARIMA expected coverage** | **Delta** | **95% CI (low)** | **95% CI (high)** | ***t* statistic** | ***p* value** |
| --- | --- | --- | --- | --- | --- | --- | --- | --- |
| **Low income** | 23 | 84.2% | 88.1% | -3.9% | -5.9% | -1.9% | - 3.83 | 1.82E-04 |
| **Lower-middle income** | 43 | 86.2% | 91.1% | -4.9% | -6.4% | -3.5% | - 6.63 | 4.75E-10 |
| **Upper-middle income** | 45 | 92.1% | 94.4% | -2.3% | -3.8% | -0.9% | - 3.20 | 0.002 |
| **High income** | 56 | 96.7% | 97.5% | -0.8% | -2.1% | 0.5% | - 1.27 | 0.21 |

(ii) *DTP3*

| **Group** | **Sample size** | **WUENIC reported coverage** | **ARIMA expected coverage** | **Delta** | **95% CI (low)** | **95% CI (high)** | ***t* statistic** | ***p* value** |
| --- | --- | --- | --- | --- | --- | --- | --- | --- |
| **Low income** | 21 | 76.2% | 81.2% | -5.0% | -7.7% | -2.3% | - 3.64 | 3.66E-04 |
| **Lower-middle income** | 45 | 81.5% | 87.0% | -5.5% | -7.3% | -3.6% | - 5.86 | 2.51E-08 |
| **Upper-middle income** | 42 | 87.3% | 91.1% | -3.8% | -5.7% | -1.9% | - 3.91 | 1.37E-04 |
| **High income** | 56 | 93.6% | 95.1% | -1.5% | -3.1% | 0.2% | - 1.77 | 0.08 |

(iii) *MCV1*

| **Group** | **Sample size** | **WUENIC reported coverage** | **ARIMA expected coverage** | **Delta** | **95% CI (low)** | **95% CI (high)** | ***t* statistic** | ***p* value** |
| --- | --- | --- | --- | --- | --- | --- | --- | --- |
| **Low income** | 21 | 75.1% | 78.9% | -3.7% | -6.5% | -0.9% | - 2.64 | 0.01 |
| **Lower-middle income** | 42 | 81.3% | 87.0% | -5.8% | -7.7% | -3.8% | - 5.77 | 4.15E-08 |
| **Upper-middle income** | 44 | 85.9% | 90.8% | -4.8% | -6.8% | -2.9% | - 4.97 | 1.75E-06 |
| **High income** | 53 | 93.4% | 94.7% | -1.3% | -3.1% | 0.4% | - 1.49 | 0.14 |

**Section S7: Sensitivity analyses for 2021**

*S7(a) T-test for all countries – sensitivity analysis, 2021*

| **Vaccine** | **Sample size** | **WUENIC reported coverage** | **ARIMA expected coverage** | **Delta** | **95% CI (low)** | **95% CI (high)** | ***t* statistic** | ***p* value** |
| --- | --- | --- | --- | --- | --- | --- | --- | --- |
| **DTP1** | 190 | 89.7% | 92.9% | -3.2% | -4.3% | -2.2% | - 6.04 | 8.17E-09 |
| **DTP3** | 190 | 84.8% | 88.6% | -3.8% | -5.1% | -2.6% | - 6.11 | 5.52E-09 |
| **MCV1** | 190 | 83.5% | 88.1% | -4.6% | -5.8% | -3.4% | - 7.39 | 4.52E-12 |

*S7(b) T-test by region – sensitivity analyses, 2021*

(i) *DTP1*

| **Group** | **Sample size** | **WUENIC reported coverage** | **ARIMA expected coverage** | **Delta** | **95% CI (low)** | **95% CI (high)** | ***t* statistic** | ***p* value** |
| --- | --- | --- | --- | --- | --- | --- | --- | --- |
| **Africa** | 53 | 84.9% | 88.2% | -3.2% | -5.2% | -1.3% | - 3.25 | 0.001 |
| **Americas** | 35 | 87.7% | 92.9% | -5.2% | -7.6% | -2.8% | - 4.24 | 3.50E-05 |
| **Asia** | 47 | 90.5% | 95.1% | -4.6% | -6.7% | -2.5% | - 4.38 | 2.00E-05 |
| **Europe** | 41 | 96.1% | 97.0% | -0.9% | -3.1% | 1.4% | - 0.76 | 0.45 |
| **Oceania** | 14 | 91.3% | 91.8% | -0.5% | -4.4% | 3.3% | - 0.28 | 0.78 |

(ii) *DTP3*

| **Group** | **Sample size** | **WUENIC reported coverage** | **ARIMA expected coverage** | **Delta** | **95% CI (low)** | **95% CI (high)** | ***t* statistic** | ***p* value** |
| --- | --- | --- | --- | --- | --- | --- | --- | --- |
| **Africa** | 53 | 78.5% | 82.0% | -3.5% | -5.8% | -1.2% | - 2.97 | 3.34E-03 |
| **Americas** | 35 | 82.1% | 88.1% | -6.0% | -8.9% | -3.2% | - 4.17 | 4.74E-05 |
| **Asia** | 47 | 87.0% | 92.7% | -5.7% | -8.2% | -3.3% | - 4.61 | 7.48E-06 |
| **Europe** | 41 | 92.7% | 94.0% | -1.3% | -3.9% | 1.4% | - 0.94 | 3.48E-01 |
| **Oceania** | 14 | 84.6% | 85.6% | -1.0% | -5.5% | 3.5% | - 0.44 | 6.63E-01 |

(iii) *MCV1*

| **Group** | **Sample size** | **WUENIC reported coverage** | **ARIMA expected coverage** | **Delta** | **95% CI (low)** | **95% CI (high)** | ***t* statistic** | ***p* value** |
| --- | --- | --- | --- | --- | --- | --- | --- | --- |
| **Africa** | 53 | 74.9% | 79.1% | -4.2% | -6.5% | -1.9% | - 3.61 | 3.95E-04 |
| **Americas** | 35 | 83.3% | 90.9% | -7.5% | -10.4% | -4.7% | - 5.24 | 4.37E-07 |
| **Asia** | 47 | 87.7% | 92.7% | -5.0% | -7.4% | -2.5% | - 4.00 | 9.22E-05 |
| **Europe** | 41 | 91.3% | 93.4% | -2.1% | -4.7% | 0.5% | - 1.57 | 0.12 |
| **Oceania** | 14 | 79.2% | 84.2% | -5.0% | -9.5% | -0.5% | - 2.20 | 0.03 |

*S7(c) T-test by income group – sensitivity analyses, 2021*

(i) *DTP1*

| **Group** | **Sample size** | **WUENIC reported coverage** | **ARIMA expected coverage** | **Delta** | **95% CI (low)** | **95% CI (high)** | ***t* statistic** | ***p* value** |
| --- | --- | --- | --- | --- | --- | --- | --- | --- |
| **Low income** | 27 | 79.5% | 85.1% | -5.6% | -8.3% | -2.9% | - 4.05 | 7.55E-05 |
| **Lower-middle income** | 53 | 86.1% | 91.1% | -5.0% | -6.9% | -3.0% | - 5.07 | 9.78E-07 |
| **Upper-middle income** | 51 | 91.4% | 94.0% | -2.5% | -4.5% | -0.5% | - 2.52 | 0.01 |
| **High income** | 58 | 96.5% | 97.4% | -0.9% | -2.8% | 0.9% | - 1.00 | 0.32 |

(ii) *DTP3*

| **Group** | **Sample size** | **WUENIC reported coverage** | **ARIMA expected coverage** | **Delta** | **95% CI (low)** | **95% CI (high)** | ***t* statistic** | ***p* value** |
| --- | --- | --- | --- | --- | --- | --- | --- | --- |
| **Low income** | 27 | 71.8% | 77.7% | -6.0% | -9.2% | -2.7% | - 3.64 | 3.51E-04 |
| **Lower-middle income** | 53 | 80.4% | 85.9% | -5.5% | -7.8% | -3.2% | - 4.69 | 5.28E-06 |
| **Upper-middle income** | 51 | 86.7% | 90.2% | -3.5% | -5.8% | -1.1% | - 2.93 | 3.78E-03 |
| **High income** | 58 | 93.7% | 95.1% | -1.4% | -3.6% | 0.8% | - 1.27 | 2.04E-01 |

(iii) *MCV1*

| **Group** | **Sample size** | **WUENIC reported coverage** | **ARIMA expected coverage** | **Delta** | **95% CI (low)** | **95% CI (high)** | ***t* statistic** | ***p* value** |
| --- | --- | --- | --- | --- | --- | --- | --- | --- |
| **Low income** | 27 | 69.3% | 75.6% | -6.2 | -9.4 | -3.1 | - 3.87 | 1.49E-04 |
| **Lower-middle income** | 53 | 78.4% | 85.1% | -6.7 | -9.0 | -4.5 | - 5.86 | 2.08E-08 |
| **Upper-middle income** | 51 | 85.5% | 90.4% | -4.8 | -7.2 | -2.5 | - 4.15 | 5.08E-05 |
| **High income** | 58 | 93.2% | 94.7% | -1.5 | -3.6 | 0.7% | - 1.35 | 0.18 |

**Section S8: Reproduced results for 2020**

Please see Section S5 for country-level reproduced results for 2020, and below global analysis. Income group, regional, and sensitivity analyses can be recreated using the R script and replacing 2021 datasets with 2020 data.

| **Vaccine** | **Sample size** | **WUENIC reported coverage** | **ARIMA expected coverage** | **Delta** | **95% CI (low)** | **95% CI (high)** | ***t* statistic** | ***p* value** |
| --- | --- | --- | --- | --- | --- | --- | --- | --- |
| **DTP1** | 168 | 90.9% | 93.7% | -2.8% | -3.6% | -2.0% | - 6.97 | 7.07E-11 |
| **DTP3** | 164 | 86.5% | 90.1% | -3.6% | -4.6% | -2.6% | - 7.20 | 2.11E-11 |
| **MCV1** | 160 | 85.8% | 89.5% | -3.8% | -4.8% | -2.7% | - 7.16 | 2.83E-11 |

**Section S9: Availability and reproducibility**

All analyses were conducted using R 4.1.2 [12]. All the data and R code implementing the analyses are publicly available from the following github repository: <https://github.com/bevans249/modelling_covid_impact_RI>

[12]	R Core Team, “R: A Language and Environment for Statistical Computing,” Vienna, Austria, 2021.
